# In a Free-Living Study of Temperature-Controlled Sleep Surfaces, Overnight Heart Rate Decreased and Heart Rate Variability Increased

**DOI:** 10.64898/2026.08.31.26361834

**Authors:** Logan Tucker, Jonathan Zwiebel, Emily Costantino, Thomas Locascio

## Abstract

**Objectives:** Temperature-controlled sleep surfaces are used nightly by hundreds of thousands of people, yet cohort analyses show improved perceived sleep quality without matching objective change. Cohort means may mask benefits concentrated in those with the greatest baseline deficit. We asked whether objective changes in sleep and overnight cardiovascular function differ by baseline sleep performance.

**Methods:** In this exploratory, retrospective, open-label study of 171 adults, wearable-derived sleep and overnight cardiovascular endpoints were compared between seven baseline nights and the first seven nights on a temperature-controlled sleep surface (Orion Sleep System). Participants were stratified by baseline performance separately for each endpoint.

**Results:** At the cohort level, total sleep time rose 7.3 minutes (p = 0.01, not significant), average heart rate fell 1.79 bpm (95% CI −2.25 to −1.34) and minimum heart rate 1.75 bpm (95% CI −2.18 to −1.33; both p < 0.001), and HRV rose 3.25 ms (95% CI 1.33 to 5.17, p = 0.001). Effects were baseline-dependent. In the most-deficient quartile for each endpoint, total sleep time rose 33.5 minutes (9.6%; 95% CI 21.5 to 45.5), REM 13.6 minutes (19.4%; 95% CI 8.6 to 18.6), deep sleep 4.4 minutes (13.1%; 95% CI 2.0 to 6.9), and HRV 4.7 ms (15.6%; 95% CI 2.2 to 7.2); wake after sleep onset fell 16.3 minutes (26.0%; 95% CI −23.0 to −9.6) in the most fragmented quartile (all p < 0.001).

**Conclusion:** Objective gains were largest where the baseline deficit was greatest.

**What Was Known:** Users of temperature-controlled sleep surfaces report clear improvements in sleep quality, but objective measurement has not confirmed them. Existing evaluations analyze participants as one group, even though sleep deficiency takes distinct forms: short sleep, fragmented sleep, and reduced deep or REM sleep. No study has asked whether these devices affect people differently depending on which deficit they carry.

**What This Study Adds:** Use of a temperature-controlled sleep surface is associated with measurable overnight cardiovascular change: heart rate fell across the cohort and in nearly every baseline quartile, and heart rate variability rose across the cohort and in those with the lowest baseline. Objective sleep benefits are not absent from these devices but are concentrated by deficit, with gains in deep sleep, REM sleep, total sleep time, and sleep continuity appearing in the participants who lacked each specific measure at baseline, while remaining near zero cohort-wide. These findings show that the long-reported gap between subjective and objective results reflects, at least in part, the averaging of heterogeneous sleepers rather than the absence of an effect.

## Introduction

Sleep is now widely recognized as a modifiable determinant of cardiometabolic, cognitive, and immune health^1–3^, and behavioral changes can improve it^4^. Thermoregulation is among the most tractable physiological levers available for altering it^5–6^. Consumer interest in modifying sleep with technology is substantial: roughly one in three U.S. adults now track their sleep digitally ^7–8^. Specifically, water-circulating covers and pads that actively heat and cool the sleep surface give users fine-grained control of surface temperature and the ability to schedule changes across the night at their own discretion^9–11^. Laboratory studies using thermosuits and specialized mattresses suggest that skin and surface temperature can influence sleep depth and heart rate, but these were brief, researcher-controlled manipulations; for consumer devices that users program themselves at home, neither an objective effect nor an effective dose has been established^12–13^. Consumer devices of this kind were a niche product until the late 2010s; they are now used nightly in hundreds of thousands of homes internationally, supplied by several competing systems and accompanied by consistently positive consumer reporting^14–15^.

Research into these devices has not scaled in proportion to adoption^16–17^. What exists divides along a single line: self-reported benefit is large and consistent, objectively measured benefit is neither^9–11^. In a randomized, counterbalanced crossover of 34 healthy adults, perceived sleep quality improved with a large effect (d = 0.92) and every daily perceptual outcome favored the device, while no objective sleep or biometric measure differed significantly^9^.

In a one-week free-living evaluation of a temperature-controlled cover (N=54, 27 male, 27 female), five of the six Pittsburgh Sleep Quality Index (PSQI)^18^ components assessed improved (all p < 0.05), while objective results were narrower: total sleep time and sleep efficiency were unchanged in both sexes, and sleep stage improvements reached significance only once filtered by sex, time of night, and temperature setting^10^. Although laboratory studies support a thermoregulatory mechanism by which surface temperature can influence sleep^5, 12–13^, whether consumer temperature-controlled surfaces produce measurable changes in sleep architecture, continuity, or autonomic function under real-world conditions has not been established. Given the number of people who now sleep on these systems, resolving whether they alter these outcomes, or whether their reception reflects thermal comfort and expectancy^19^, is a critical and unmet priority. This exploratory analysis addresses that question in a free-living cohort, examining each endpoint separately by baseline performance rather than as a single cohort average; no hypotheses were specified in advance, and the quartile-level findings are reported as hypothesis-generating.

## Study Overview

A total of 171 adults (mean age 40.3 ± 11.3 years; 121 male, 50 female) contributed overnight physiological data via Apple Watch spanning their transition onto the Orion Sleep System between January and July 2026, yielding 2,394 participant-nights: seven baseline nights and seven on-device nights for every participant. Figure 1 shows the sleep surface, the wearable, the staging model that converts its sensor streams into 30-second epoch labels, meaning the classifier assigns a single sleep stage to every fixed 30-second window of the recording, and the app through which participants set their own nightly temperature schedules. The analyzable sample size varies by endpoint according to sensor availability on the wearable rather than participant dropout: 171 participants for total sleep time and wake after sleep onset, 170 for deep and REM sleep, 158 for overnight heart rate, and 150 for heart rate variability. For each endpoint, participants were ranked on their baseline value alone and divided into quartiles, so that Q1 denotes the 25% with the greatest baseline deficit on that endpoint and Q4 the 25% with the least; quartile membership therefore differs across endpoints.

**Figure 1.**
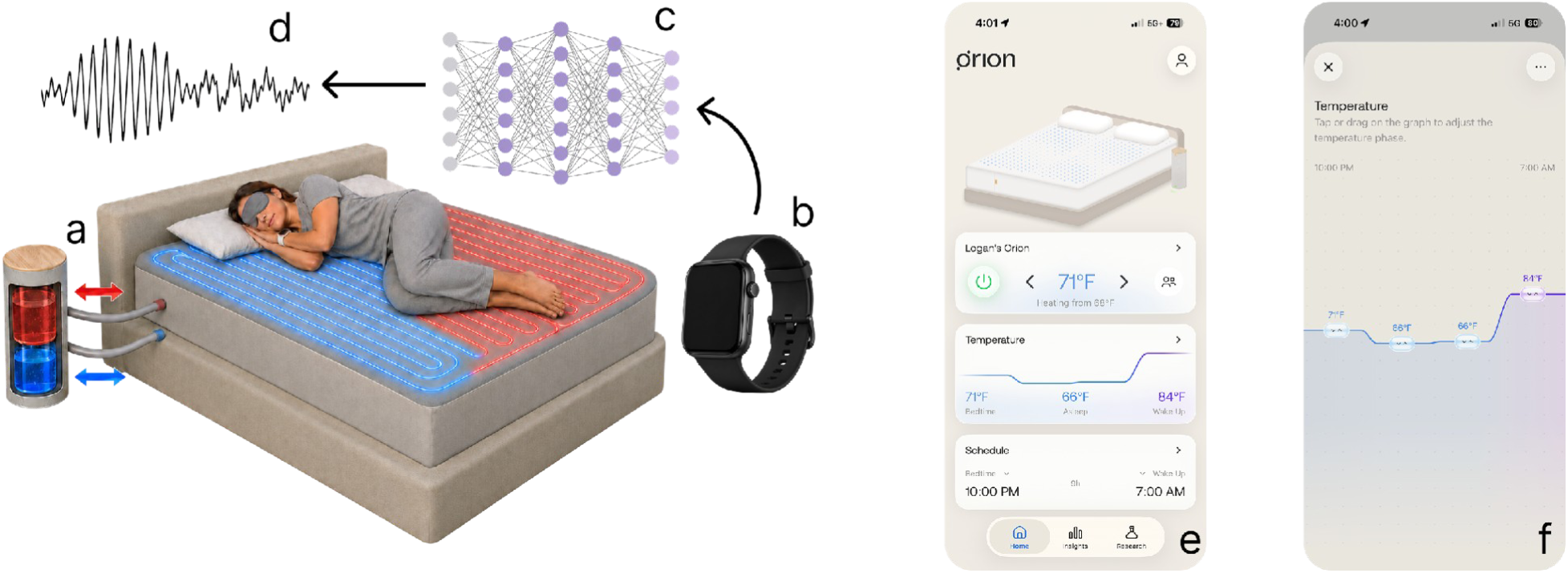
The intervention, the measurement, and the participant-facing controls. (a) The Orion Sleep System, which sets the temperature of the sleep surface by circulating temperature-treated water through channels beneath the sleeper. (b) The wrist-worn consumer device that every participant wore overnight and throughout the day. (c) The sleep staging model, which takes the device sensor streams as its inputs. (d) The model output across the night, one of wake, light sleep, deep sleep, or REM for every 30-second epoch. (e) and (f) The companion app for controlling the sleep system’s surface temperature. Participants set target temperatures at successive time-anchored set points, so temperature varied across the night. (e) Home screen showing regulation status (heating/cooling, target, current temperature) and on/off times. (f) Temperature-profile editor with set points adjusted by tapping or dragging.

## Materials and Methods

### Study Design and Participants

This retrospective, open-label, within-subject study compared sleep and overnight cardiovascular endpoints before and after participants began using an actively thermoregulated sleep surface. Data were drawn from users who, during product onboarding, opted in to sync their wearable data with the Orion app and consented to the use of their de-identified data in research with academic and other partners; users could decline and continue using their product without this feature. Participants were not recruited or instructed; this is an analysis of natural, free-living engagement with the sleep system and the wearable. The analysis plan was developed only after all data had accrued. Records were stripped of direct identifiers and assigned study identifiers before any analysis was performed, and the investigators had no interaction with the individuals whose data were analyzed and no access to identifiable private information. The work, therefore, does not meet the definition of human subjects research under 45 CFR 46.102(e), and it was not submitted for institutional review board review.

### Wearable Measurement

All participants wore an Apple Watch (Apple Inc., Cupertino, CA, USA) during sleep, and all physiological data were obtained through HealthKit. Within detected sleep, heart rate was sampled at a median inter-sample interval of 4.3 min (IQR 2.2 to 5.6 min; median 94 samples per night), respiratory rate at 10.0 min, and heart rate variability (SDNN) approximately four times per night^20^. Sleep staging is a validated model output rather than a sensor reading. The classifier draws on the full sensor complement of the device, comprising a three-axis accelerometer, a gyroscope, an optical heart sensor, an electrical heart sensor, a blood oxygen sensor, a wrist temperature sensor, and an ambient light sensor, and returns variable-length stage runs quantized to a 30 s grid as light, deep, REM, or awake.

### Intervention and Open-Label Temperature Use

The Orion Sleep System is a cover that fits over any existing mattress. Soft water channels, imperceptible to the sleeper, run throughout the sleep surface and connect to a bedside controller that treats the circulating water with thermoelectric modules, holding it at a commanded temperature with fine granularity in either direction. Because the water flows immediately beneath the sleeper, the commanded water temperature sets the temperature of the sleep surface itself, so the device acts directly on the sleeper’s thermal environment rather than on the temperature of the room.

The device is open-label by design. A companion mobile application gives the participant direct control of that temperature and of how it moves across the night, structured as four set points: a bedtime temperature the bed is preconditioned to before the participant gets in, a night phase a few hours later, a dawn phase a few hours before waking, and a wake-up temperature (Figure 1e, f). Each is set independently, so a participant can, for example, run the bed cool early, warm in the middle of the night, and cool again toward waking, and can change that pattern from night to night. No sham, blinding, or assigned temperature protocol was used. Commanded set points were recoverable for 1,077 of the 1,197 on-device nights (90.0%, 158 of 171 participants). The mean was 22.3 °C (72.0 °F), but participant means spanned 10.8 to 32.3 °C (SD 4.01 °C), and 73% of nights warmed from the first half to the second, the cohort mean rising from 20.9 °C at bedtime to 24.8 °C at wake-up.

### Baseline and Treatment Windows

For each participant, the baseline window comprised the seven most recent nights with usable recordings preceding their first night on the device, and the treatment window comprised their first seven nights with usable recordings on the device. A night was unusable if the participant did not wear the wearable to bed, or, within the treatment window, did not sleep on the device. Nightly values were averaged within each window, producing one baseline and one on-device value per participant per endpoint. Because data were collected in free-living conditions, the seven nights in each window were not necessarily consecutive. The seven on-device nights accrued over a median of 8 calendar nights (IQR 7 to 11), and the seven baseline nights required a median of 7 nights (IQR 7 to 9). Uninterrupted windows were supplied by 36.8% of participants on the treatment side and 54.4% on the baseline side, and for 86.5%, the final baseline night immediately preceded device onset.

### Endpoints and Baseline Stratification

Endpoints were total sleep time, wake after sleep onset, deep sleep in minutes and as a percentage of total sleep time, REM sleep in minutes and as a percentage, minimum and average overnight heart rate, and heart rate variability. Analyzable N varies by endpoint according to sensor availability: 171 for continuity measures, 170 for sleep architecture, 158 for heart rate, and 150 for heart rate variability. For each endpoint, participants were ranked on their baseline value alone and split into four equal groups. Q1 is the 25% of participants with the worst baseline value on that endpoint, and Q4 the 25% with the best. Each participant’s baseline is then compared with their own on-device value. Membership differs across endpoints, since a participant may fall in Q1 for one endpoint and Q4 for another.

### Statistical Analysis

Cohort-level and quartile-level comparisons used two-sided paired t-tests on participant-level means, with 95% confidence intervals reported for each mean difference. Because the dependent variables were highly correlated and some quartile groups were small, the assumptions for an omnibus repeated-measures MANOVA were often not met, so paired t-tests were used for all comparisons. To account for multiple comparisons, statistical significance was set at p < 0.005, based on the Hochberg step-up multiplicity adjustment procedure; unadjusted p-values are reported throughout. Stratification on baseline values makes regression toward the mean an expected contributor to quartile-level effects. All analyses were performed by an independent biostatistician in R (R Foundation for Statistical Computing, Vienna, Austria).

## Results

### Overnight Cardiovascular Function

Average heart rate and minimum heart rate fell during the first week on the device (Table 1). Across the cohort, average overnight heart rate declined 1.79 bpm (64.1 to 62.3 bpm, 95% CI −2.25 to −1.34, p < 0.001) and minimum overnight heart rate declined 1.75 bpm (57.9 to 56.2 bpm, 95% CI −2.18 to −1.33, p < 0.001). These were the most uniform changes observed in the study: every quartile moved in the same direction, and seven of eight quartile-level tests reached significance, so the change was not confined to participants with elevated baseline rates. The magnitude, however, was graded by baseline: minimum heart rate fell 2.8 bpm in Q1 against 1.1 bpm in Q4, and average heart rate fell 2.6 bpm in each of the two most-elevated quartiles against 0.7 bpm in Q4, a change that did not reach significance (95% CI −1.40 to −0.03, p = 0.04).

**Table 1.** Overnight cardiac endpoints before and during the first week on the Orion Sleep System, by baseline quartile. Quartiles are ranked on baseline values, so Q1 is the greatest-need group for each endpoint. Change is the percentage change in the group mean; the 95% confidence interval is on the raw within-participant difference in the units of the endpoint. p-values below 0.005 are set in bold.

| Group | n | Before | On Orion | Change | 95% CI | p |
| --- | --- | --- | --- | --- | --- | --- |
| **Minimum heart rate (bpm)** |  |  |  |  |  |  |
| **All participants** | **158** | **57.9** | **56.2** | **-3.0%** | **-2.18 to -1.33** | **< 0.001** |
| Q1 (greatest need) | 40 | 68.7 | 65.9 | -4.1% | -3.81 to -1.85 | < 0.001 |
| Q2 | 40 | 60.0 | 58.2 | -3.1% | -2.88 to -0.79 | **0.001** |
| Q3 | 39 | 54.4 | 53.1 | -2.3% | -2.00 to -0.51 | **0.002** |
| Q4 (least need) | 39 | 48.3 | 47.2 | -2.2% | -1.61 to -0.51 | < 0.001 |
| **Average heart rate (bpm)** |  |  |  |  |  |  |
| **All participants** | **158** | **64.1** | **62.3** | **-2.8%** | **-2.25 to -1.34** | **< 0.001** |
| Q1 (greatest need) | 40 | 75.7 | 73.1 | -3.4% | -3.56 to -1.67 | < 0.001 |
| Q2 | 40 | 66.6 | 64.0 | -3.9% | -3.77 to -1.47 | < 0.001 |
| Q3 | 39 | 60.2 | 59.0 | -2.0% | -1.88 to -0.47 | 0.002 |
| Q4 (least need) | 39 | 53.4 | 52.7 | -1.3% | -1.40 to -0.03 | 0.042 |
| **Heart rate variability (ms)** |  |  |  |  |  |  |
| **All participants** | **150** | **50.6** | **53.8** | **+6.4%** | **1.33 to 5.17** | **0.001** |
| Q1 (greatest need) | 38 | 30.1 | 34.8 | +15.6% | 2.24 to 7.17 | < 0.001 |
| Q2 | 38 | 41.0 | 44.1 | +7.6% | 0.49 to 5.71 | 0.021 |
| Q3 | 37 | 52.5 | 54.1 | +3.1% | -2.72 to 5.93 | 0.457 |
| Q4 (least need) | 37 | 79.6 | 83.1 | +4.5% | -2.13 to 9.25 | 0.213 |

Figure 2 shows the change for each participant individually. Average overnight heart rate fell in 113 of 158 participants (71.5%) and minimum heart rate in 118 of 158 (74.7%), with the two panels drawn on a shared vertical axis. The cohort-level declines therefore reflect a shift across most of the distribution rather than a large movement in a minority of participants.

**Figure 2.**
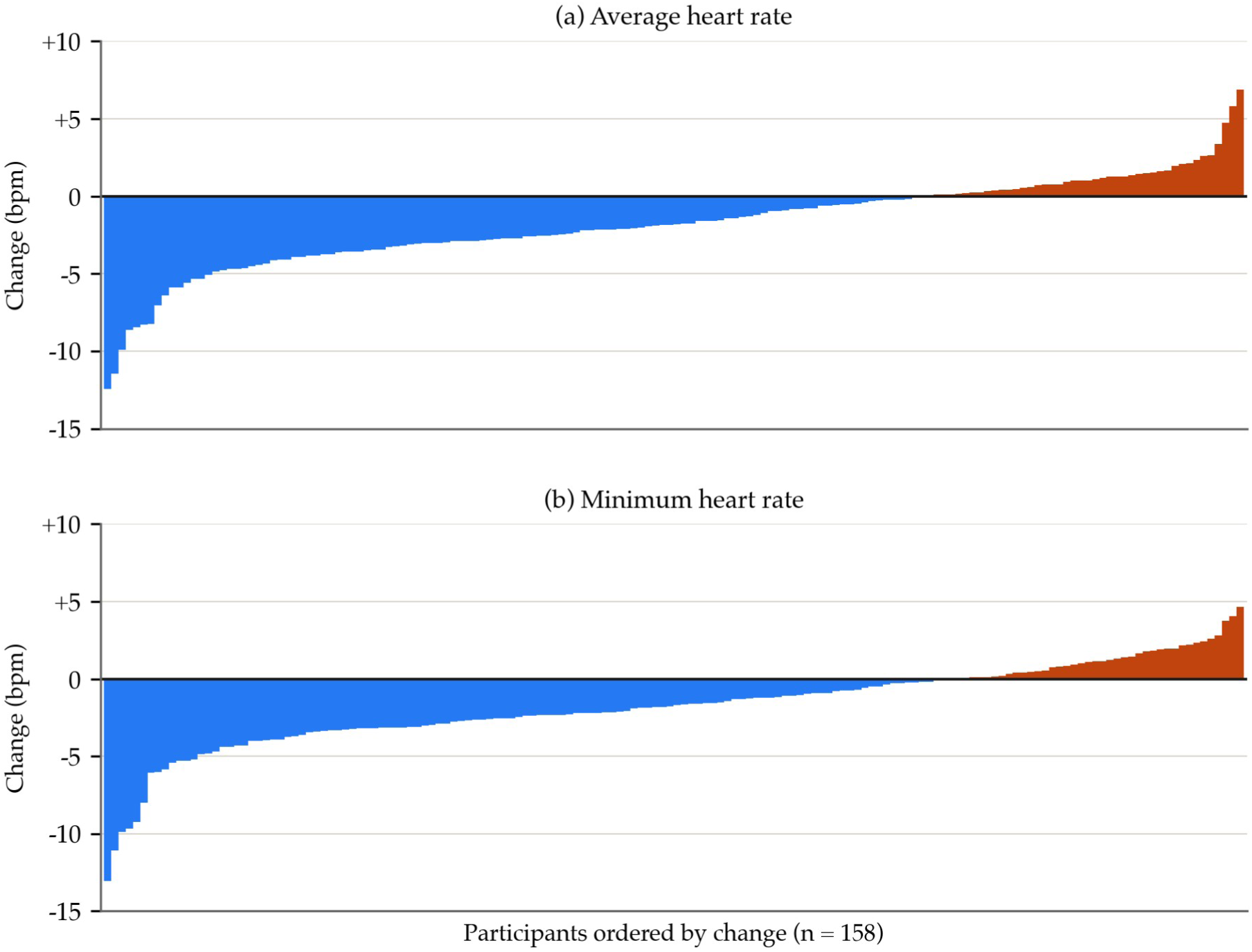
Change in overnight heart rate for every participant with heart rate data in both windows, one bar per participant, ordered from the largest fall to the largest rise. (a) Average overnight heart rate. (b) Minimum overnight heart rate. Each participant is summarized by the mean of their nights within each window, and the change is their on-Orion mean minus their baseline mean. Bars below the zero line are participants whose heart rate fell, bars above it those whose rate rose. Both panels share the same vertical axis, so the two distributions can be compared directly.

Heart rate variability rose 3.25 ms across the cohort (50.6 to 53.8 ms, 95% CI 1.33 to 5.17, p = 0.001), but the quartile pattern differed from that for heart rate. The gain reached significance only in participants with the lowest baseline HRV: 4.7 ms in Q1 (30.1 to 34.8 ms, 95% CI 2.24 to 7.17, p < 0.001). Q2 rose 3.1 ms (95% CI 0.49 to 5.71, p = 0.02), and Q3 (p = 0.46) and Q4 (p = 0.21) also rose, but none of these three changes reached significance.

### Total Sleep Time and Sleep Continuity

Total sleep time increased across the cohort, from 410.3 to 417.6 minutes, a change that did not reach significance (95% CI 1.74 to 12.84, p = 0.01; Table 2). The magnitude, again, varied by baseline. The gain was largest in the participants who slept least at baseline. The lowest quartile gained 33.5 minutes (347.6 to 381.1 minutes, a 9.6% increase, 95% CI 21.47 to 45.52, p < 0.001). Q2 gained 8.9 minutes (95% CI −2.00 to 19.86, p = 0.11), and Q3 gained 4.3 minutes (95% CI −3.82 to 12.49, p = 0.29), neither reaching significance, consistent with participants already sleeping adequately having less room to gain. Total sleep time declined 18.2 minutes in Q4, the quarter sleeping longest at baseline (473.2 to 455.0 minutes, 95% CI −26.00 to −10.37, p < 0.001).

**Table 2.** Total sleep time and wake after sleep onset before and during the first week on the Orion Sleep System, by baseline quartile. Quartiles are ranked on baseline values (Q1 is the greatest-need group for each endpoint). Change is the percentage change in the group mean; the 95% confidence interval is on the raw within-participant difference in minutes. p-values below 0.005 are set in bold.

| Group | n | Before | On Orion | Change | 95% CI | p |
| --- | --- | --- | --- | --- | --- | --- |
| **Total sleep time (min)** |  |  |  |  |  |  |
| All participants | 171 | 410.3 | 417.6 | +1.8% | 1.74 to 12.84 | 0.010 |
| Q1 (greatest need) | 43 | 347.6 | 381.1 | +9.6% | 21.47 to 45.52 | **< 0.001** |
| Q2 | 43 | 395.5 | 404.4 | +2.3% | -2.00 to 19.86 | 0.107 |
| Q3 | 43 | 426.5 | 430.9 | +1.0% | -3.82 to 12.49 | 0.290 |
| Q4 (least need) | 42 | 473.2 | 455.0 | -3.8% | -26.00 to -10.37 | **< 0.001** |
| **Wake after sleep onset (min)** |  |  |  |  |  |  |
| All participants | 171 | 33.3 | 31.6 | -5.1% | -4.47 to 1.06 | 0.226 |
| Q1 (greatest need) | 43 | 62.7 | 46.4 | -26.0% | -23.04 to -9.63 | **< 0.001** |
| Q2 | 43 | 36.6 | 37.4 | +2.2% | -5.01 to 6.61 | 0.782 |
| Q3 | 43 | 22.0 | 25.4 | +15.8% | 0.41 to 6.53 | 0.027 |
| Q4 (least need) | 42 | 11.5 | 16.9 | +47.2% | 1.90 to 8.93 | **0.003** |

Cohort-wide, wake after sleep onset fell 5.1%, from 33.3 to 31.6 minutes, a change that did not reach significance (95% CI −4.47 to 1.06, p = 0.23).

The cohort figure holds one large exception. Among participants with the most fragmented sleep at baseline, wake time fell 16.3 minutes, from 62.7 to 46.4 minutes, a 26.0% reduction (95% CI −23.04 to −9.63, p < 0.001). This was the only quartile in which fragmentation fell, and its decline was roughly three times the magnitude of the largest change recorded in any other stratum. The participants it reaches are those whose baseline fragmentation was severe, averaging more than an hour awake after sleep onset each night, and this pattern is consistent with an effect on sleep maintenance specifically, rather than on sleep continuity in general.

Figure 3 renders this relationship at the level of the individual participant. It shows the correspondence between how much support a participant needed and how much benefit they received. The largest reductions in wake time are concentrated among the participants who entered the study with the greatest need, and the benefit diminishes across the ordered cohort as baseline need falls.

**Figure 3.**
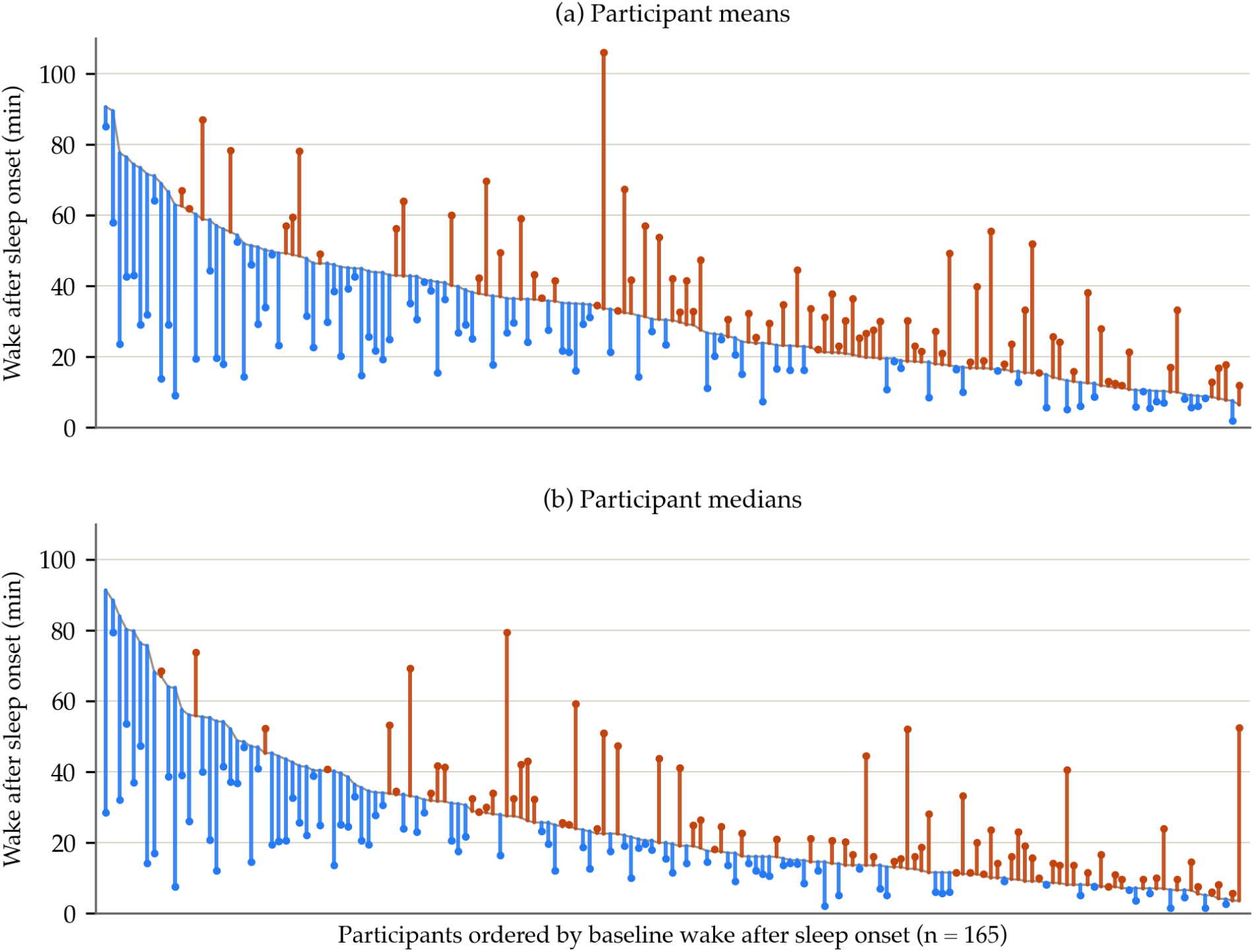
Wake after sleep onset for every participant, before Orion and during their first week on it, summarized two ways. Ordered from the greatest baseline need at the left, where fragmentation was most severe, to the least at the right, and drawn twice to summarize each participant’s nights by mean and by median. (a) Each participant’s mean across the nights in each window. (b) Each participant’s median across the same nights. One stem per participant, running from their baseline value to their on-Orion value, with the marker at the on-Orion end; blue stems are participants whose wake time fell, while orange stems those whose wake time rose. The gray trace is the sorted baseline. Each panel is ordered independently on its own baseline statistic, highest baseline at the left, therefore a participant’s position can differ between panels. Six of the 171 participants are not shown: the 3 with the highest baseline wake time and the 3 with the lowest, the 2% at each end of the baseline distribution, leaving 165 plotted with baselines from 6.3 to 90.6 min.

### Sleep Architecture: Deep and REM Sleep

Neither architecture endpoint changed substantively across the cohort. Deep sleep rose 1.2 minutes (46.9 to 48.2 minutes, 95% CI −0.05 to 2.54, p = 0.06) and REM sleep rose 1.7 minutes (99.2 to 101.0 minutes, 95% CI −0.90 to 4.35, p = 0.20; Table 3).

**Table 3.** Deep sleep and REM sleep in minutes before and during the first week on the Orion Sleep System, by baseline quartile. Quartiles are ranked on baseline values, so Q1 is the greatest-need group for each endpoint. Change is the percentage change in the group mean; the 95% confidence interval is on the raw within-participant difference in minutes. p-values below 0.005 are set in bold. *Approached significance (0.005 < p < 0.01).

| Group | n | Before | On Orion | Change | 95% CI | p |
| --- | --- | --- | --- | --- | --- | --- |
| Deep sleep (min) |  |  |  |  |  |  |
| All participants | 170 | 46.9 | 48.2 | +2.7% | -0.05 to 2.54 | 0.060 |
| Q1 (greatest need) | 43 | 33.7 | 38.1 | +13.1% | 1.95 to 6.88 | < **0.001** |
| Q2 | 43 | 43.0 | 45.0 | +4.7% | -0.23 to 4.29 | 0.077 |
| Q3 | 42 | 49.4 | 50.0 | +1.3% | -1.66 to 2.91 | 0.583 |
| Q4 (least need) | 42 | 62.1 | 59.9 | -3.5% | -5.38 to 1.00 | 0.172 |
| REM sleep (min) |  |  |  |  |  |  |
| All participants | 170 | 99.2 | 101.0 | +1.7% | -0.90 to 4.35 | 0.196 |
| Q1 (greatest need) | 43 | 70.0 | 83.6 | +19.4% | 8.61 to 18.57 | < **0.001** |
| Q2 | 43 | 92.7 | 94.9 | +2.4% | -2.44 to 6.84 | 0.344 |
| Q3 | 42 | 107.2 | 104.1 | -2.9% | -8.65 to 2.41 | 0.261 |
| Q4 (least need) | 42 | 127.9 | 121.8 | -4.7% | -10.35 to -1.74 | 0.007* |

Again, both endpoints changed in the quartile with the lowest baseline value. Deep sleep rose 4.4 minutes, from 33.7 to 38.1 minutes (95% CI 1.95 to 6.88, a 13.1% increase, p < 0.001). REM sleep rose 13.6 minutes, from 70.0 to 83.6 minutes (95% CI 8.61 to 18.57, a 19.4% increase, p < 0.001). These were the only substantive increases on either endpoint.

Elsewhere, changes were small or negative. Deep sleep did not change materially in Q2, Q3, or Q4 (p = 0.08, 0.58, and 0.17). REM sleep did not change significantly in Q2 or Q3 (p = 0.34 and 0.26), and a 6.0-minute decline in Q4, from 127.9 to 121.8 minutes, approached significance (95% CI −10.35 to −1.74, p = 0.007).

## Discussion

The largest and most consistent changes in this cohort were cardiovascular, and this is also where the measurement platform is strongest^21^. Heart rate is measured directly by the device’s optical sensor, whereas sleep stage is the output of a classifier model and inherits that model’s prediction error; the cardiac endpoints are therefore the most direct and accurate measurements in this study^22–23^. Average overnight heart rate fell 1.79 bpm (95% CI −2.25 to −1.34, p < 0.001) and minimum overnight heart rate 1.75 bpm (95% CI −2.18 to −1.33, p < 0.001), while heart rate variability rose 3.25 ms (95% CI 1.33 to 5.17, p = 0.001). This decline is similar in magnitude to the heart-rate reduction reported in independent polysomnographic work on nocturnal body cooling, and the concurrent HRV rise is consistent with a cooling-driven increase in high-frequency, vagally mediated HRV^13, 24^. Nocturnal heart rate and HRV are independent markers of cardiovascular risk^25, 26^. Both endpoints carry prognostic weight: higher nighttime heart rate independently predicts cardiovascular events (hazard ratio 1.17 per 10 bpm)^25^, and each 1% increase in SDNN is associated with roughly 1% lower cardiovascular risk^26^. These are long-term, between-person associations and do not imply a risk reduction from a one-week change, but they establish the clinical relevance of the endpoints that moved most consistently here. Perhaps more important than the cohort means is their uniformity: every cardiovascular measure moved in the favorable direction in every baseline quartile, seven of eight quartile-level heart rate tests reached significance, and these results rest on the measurements this platform performs most reliably. The convergence of three directly sensed endpoints strengthens the finding beyond the p-values alone.

Prior evaluations of these devices have reported subjective gains that objective measurement has not matched^9–11^. Sleep deficiency takes many forms, including short sleep duration, fragmented sleep with frequent nighttime awakenings, and reduced slow-wave or REM sleep^27–28^. A cohort-wide mean collapses each of these sleeper types into a single number, which does not allow the impact of these devices to be understood for any specific sleeper type. By assembling cohorts large enough to subdivide, and then stratifying them by condition, that impact can be measured. Because quartile membership is computed per endpoint, a participant may fall in the greatest-need quartile for fragmentation and the least-need quartile for duration, so each stratification assembles the individuals carrying that specific deficiency and asks what changed for them. Closing the gap between robust subjective findings and the objective measures that have not matched them will require larger cohorts, stratified by specific sleep deficit, with each deficit analyzed separately.

The sleep architecture and continuity endpoints show this pattern directly. Deep sleep rose 1.2 minutes cohort-wide and did not reach significance (p = 0.06); within the quartile with the least deep sleep at baseline, it rose 4.4 minutes, a 13.1% increase (95% CI 1.95 to 6.88, p < 0.001). Conductive body cooling has independently been shown to increase slow-wave sleep¹³, and the cohort’s mean commanded set point of 22.3 °C, well below typical skin temperature, indicates that the surface predominantly cooled sleepers in this study, consistent with the deep-sleep gain observed in the most deficient quartile. REM rose 1.7 minutes cohort-wide, also non-significant (p = 0.20); within the quartile with the least REM, it rose 13.6 minutes, a 19.4% increase (95% CI 8.61 to 18.57, p < 0.001). Wake after sleep onset fell 1.7 minutes cohort-wide (p = 0.23) and 16.3 minutes in the most fragmented quartile, a 26.0% reduction (95% CI −23.04 to −9.63, p < 0.001). This concentration among the most fragmented sleepers parallels earlier work in which cutaneous warming reduced nighttime wakefulness most in those with the greatest baseline fragmentation^12^. Total sleep time rose 7.3 minutes cohort-wide, also non-significant (95% CI 1.74 to 12.84, p = 0.01); within the shortest-sleeping quartile, it rose, significantly, 33.5 minutes (95% CI 21.47 to 45.52, p < 0.001). Read only as cohort averages, all four endpoints are null results. Read by sleeper type, each one carries a substantial and highly significant gain in the group defined by that deficiency.

What these findings call for is a randomized trial against a fixed-temperature control, enriched for the most deficient sleepers, validated against polysomnography, and run well beyond a week. The set point data collected here would also support a dose-response analysis, which this study did not attempt.

## Limitations

Several limitations bear on how these results should be read. First, stratification on baseline values makes regression to the mean an expected contributor to the quartile-level effects, because sleep varies from night to night: participants ranked lowest on a single seven-night window are more likely to have been captured during an unusually poor stretch, and their values would tend to improve regardless of intervention^29^. Quartile membership is more stable for the cardiac endpoints than for sleep architecture (Supplementary Figure 2).

Second, this single-arm pre-post design lacks a fixed-temperature comparison, so the observed changes cannot be fully separated from novelty, expectancy, seasonality, or concurrent behavior change^30^. These explanations bear most directly on endpoints a participant can influence and least on overnight heart rate, which is not under voluntary control and declined consistently across every baseline stratum. This analysis prioritized the largest feasible cohort to examine variation across sleeper types and baseline sleep deficits, rather than long-term outcomes.

Third, the device is open-label by design, with no sham, no blinding, and no assigned thermal protocol; participant mean set points varied widely, so the reported effects are averages over heterogeneous self-selected exposures rather than the effect of a single prescribed one, and expectancy cannot be excluded in a paradigm that has previously produced large perceptual effects alongside null objective ones.

In addition, HRV was sampled far less densely than heart rate, at approximately four samples per night, and the cohort-level estimate should be read with that in mind.

Finally, sleep stage is the output of a classifier rather than a direct sensor reading and was not validated against polysomnography in this cohort, and because data were captured in free-living use, which wrist the device was worn on, watch model, and band fit were neither standardized nor recorded.

## Data Availability

Data referenced in the study is not available at this time.

## Supplemental Information

### Open-Label Temperature Use

The open-label nature of the device is a critical feature of this study. Participants set their own temperatures, so each one received a thermal exposure of their own choosing rather than a common prescribed one. Supplementary Figure 1 shows how far apart those choices were, and therefore how differently the sleep surface was thermally augmented across the analyzed cohort.

**Supplementary Figure 1.**
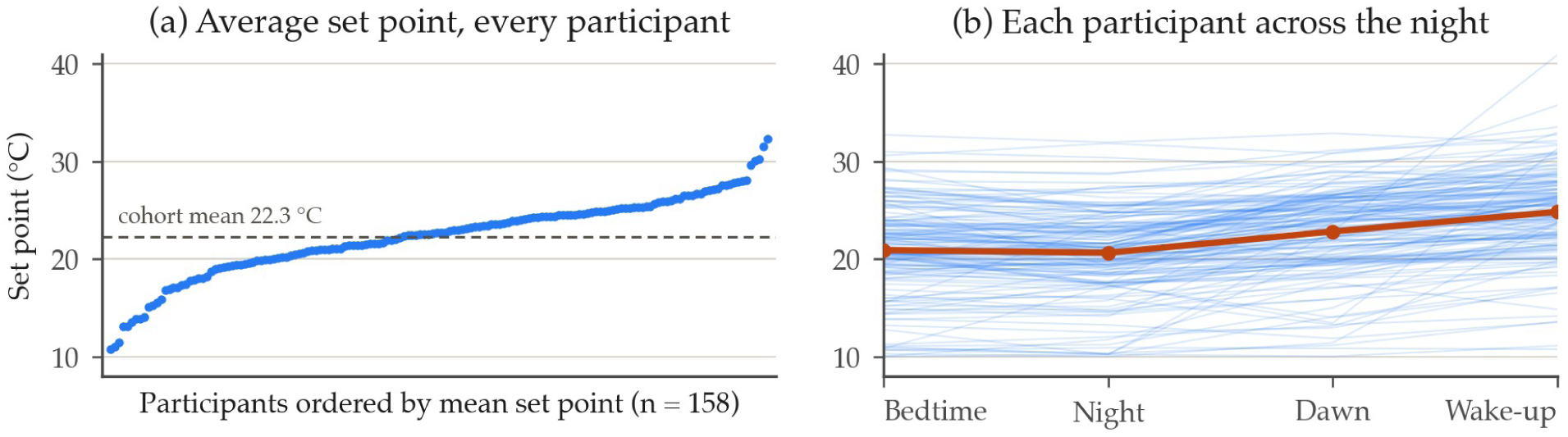
Set point use across the cohort, one point or line per participant. (a) Each participant’s average commanded set point across their nights on the device, ordered from coldest to warmest, with the cohort mean of 22.3 °C drawn dashed. (b) The same participants across the four set points of the night, each faint line one participant and the heavy line the cohort mean, which runs from 20.9 °C at bedtime to 24.8 °C at wake-up. Both panels cover the 158 participants whose on-device nights carried set point data.

### Baseline Representativeness

Ranking participants into quartiles on a seven-night baseline invites an objection: if that window caught the worst quartile on an unusually bad stretch, part of their apparent improvement is regression to their own mean.

For each participant, the seven usable nights immediately before the study baseline were scored by the same criteria, reaching no farther back than 30 days. The study baseline is the seven-night window the study ranked people on, and the preceding week is the seven usable nights directly before it. A full preceding week exists for 146 to 167 participants, varying by endpoint with sensor availability.

Ranked on the preceding week instead of the study baseline, the share of participants who land in the same quartile, and the share who land within one quartile of it, were: total sleep time 51% and 91%, wake after sleep onset 47% and 87%, deep sleep 44% and 82%, REM sleep 51% and 92%, minimum heart rate 81% and 99%, average heart rate 83% and 100%, heart rate variability 62% and 98%.

**Supplementary Figure 2.**
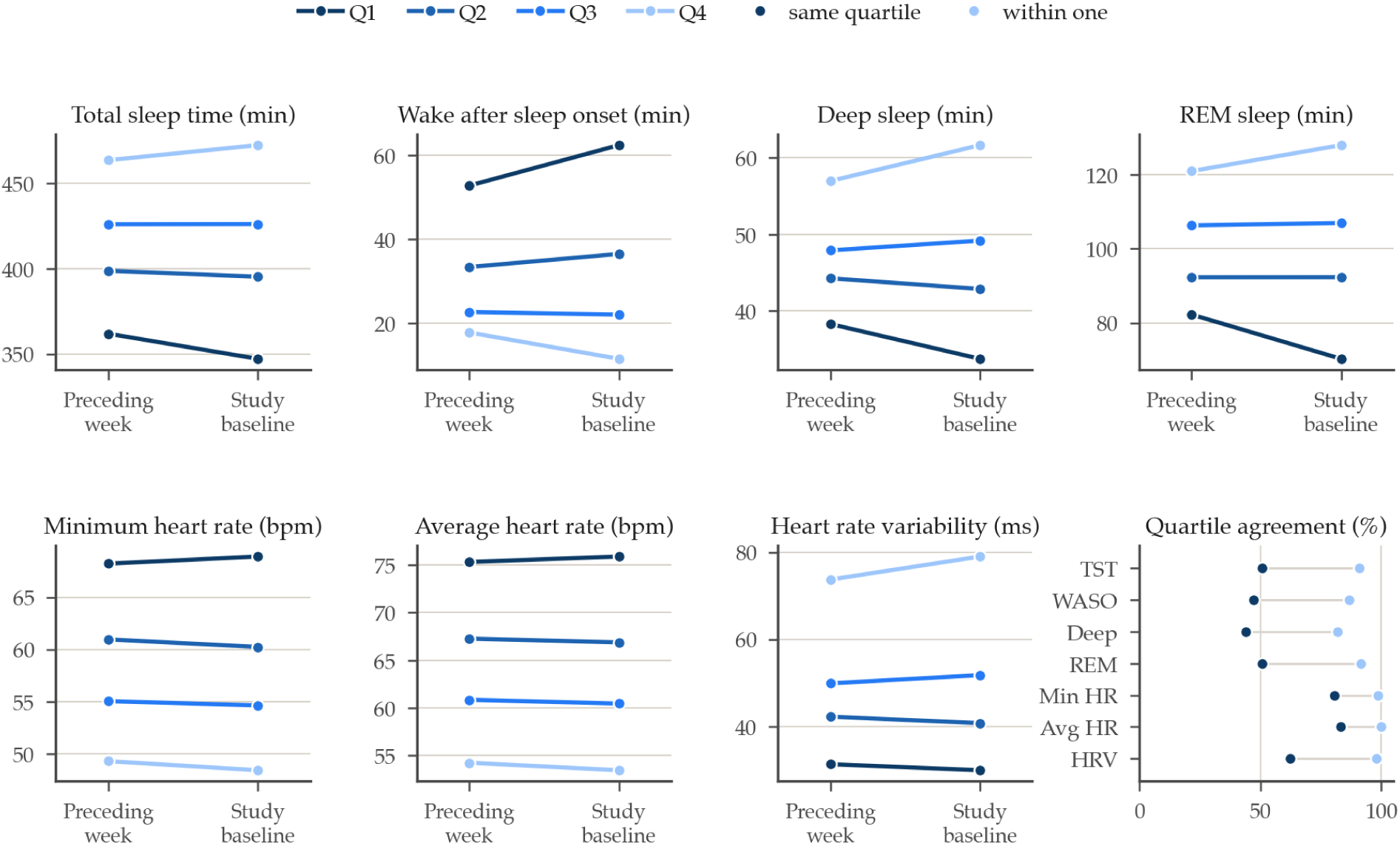
The preceding week against the study baseline. Each of the first seven panels is one endpoint in its own units, showing where each study-baseline quartile sat on the two weeks. The last panel gives the share of participants landing in the same quartile when ranked on the preceding week, and the share landing within one quartile of it. 146 to 167 participants per endpoint.

